# Individual and Population Level Uncertainty Interact to Determine the Performance of Outbreak Surveillance Systems

**DOI:** 10.1101/2025.07.11.25331369

**Authors:** Callum R.K. Arnold, Alex C. Kong, Amy K. Winter, William J. Moss, Bryan N. Patenaude, Matthew J. Ferrari

## Abstract

**Background:** Outbreak detection frequently relies on imperfect individual-level case diagnosis. Both outbreaks and cases are discrete events that can be misclassified and uncertainty at the case level may impact the performance of outbreak alert and detection systems. Here, we describe how the performance of outbreak detection depends on individual-level diagnostic test characteristics and population-level epidemiology and describe settings where imperfect individual-level tests can achieve consistent performance comparable to “perfect” diagnostic tests.

**Methodology:** We generated a stochastic SEIR model to simulate daily incidence of measles (i.e., true) and non-measles (i.e., noise) febrile rash illness. We modeled non-measles sources as either independent static (Poisson) noise, or dynamical noise consistent with an independent SEIR process (e.g., rubella). Defining outbreak alerts as the exceedance of a threshold by the 7-day rolling average of observed test positives, we optimized the threshold that maximized outbreak detection accuracy across set of noise structures and magnitudes, diagnostic test accuracy (consistent with either a perfect test, or proposed rapid diagnostic tests), and testing rates.

**Conclusions:** The optimal threshold for each diagnostic test typically increased monotonically with testing rate. With static noise, outbreak detection with RDT-like and perfect tests achieved accuracies of 90%, with comparable delays to outbreak detection. With dynamical noise, the accuracy of perfect test scenarios was superior to those achieved with RDTs (≈ 90% vs.≈ 80%). Outbreak detection accuracy declined as dynamical noise increased and leads to permanent alert status with RDT-like tests at very high noise. The performance of an outbreak detection system is highly sensitive to the structure and the magnitude of the background noise. Depending on the epidemiological context, outbreak detection using RDTs can perform as well as perfect tests.

**Author Summary:** To respond to outbreaks of infectious diseases, we first need to detect them. This detection is inherently flawed, in part, due to imperfect diagnostic tests used to indicate whether individuals are positive or negative for a disease. We evaluated the impact of imperfect diagnostic tests for infectious disease on the accuracy and timeliness of outbreak detection in the context of a set of background infections that could be mistaken for the disease of interest, and consequently cause false positive test results. We find that when outbreak detection performance is highly dependent on the structure and magnitude of the background “noise” infections. When the rate of background infections far exceeds that of the target infection, and are dynamical, such that there are large peaks and troughs of “noise infections”, imperfect diagnostic tests are not able to accurately distinguish the “signal” (target infections) from the background “noise”. If the background “noise” infections are either less cyclical in their dynamics, or do not outnumber true infections by a great deal, imperfect diagnostic tests can perform well.

## Background

Infectious disease diagnostics are medical devices and techniques that can be used to detect the presence of a pathogen in a host [1]. A clinician may use a physical examination to diagnose a patient with an infection, identifying the signs and symptoms that result from the host’s immune response to the pathogen (e.g., fever, rash). Alternatively, *in vitro* tests to quantify the presence of the pathogen itself may be used, e.g., polymerase chain reaction (PCR) to detection pathogen nucleic acids, or the host’s immune response to the pathogen e.g., enzyme-linked immunosorbent assays (ELISA) to measure IgM or IgG antibody responses [1–3]. Any given diagnostic will vary in its ability to correctly identify the presence of the pathogen, which is described by its sensitivity and specificity. The sensitivity of a diagnostic is the ability to correctly identify a positive result, conditional on a positive individual being tested i.e., a true positive result [4–6]. The specificity is the opposite: the ability to correctly determine a true negative result, conditional on a negative individual being tested [4–6]. Due to the translation of quantitative measures e.g., immunoglobulin M (IgM) antibody titers, into a binary outcome (positive/negative), the sensitivity and specificity of a diagnostic are often at odds with one another. For example, using a low optical density value to define the threshold for detection for an ELISA will produce a diagnostic that is highly sensitive, as it only requires a small host response to the pathogen and many resulting antibody titers will exceed this value. However, this may lead to low specificity due to an increase in spurious false positive results in non-infected individuals. To account for these differences, the target product profile (TPP) of a diagnostic provides a minimum set of characteristics that should be met, helping to guide the development and use [7].

The choice to prioritize sensitivity or specificity will be pathogen and context specific. When the cost of a false negative result is disproportionately high relative to a false positive, such as for Ebola [8], highly specific tests may be preferred. This balance will, however, vary as the prevalence of infection in a population varies. Higher prevalence of infection in a population will increase the positive predictive value (PPV) of the test i.e., the probability that a positive test reflects an infected individual, that unlike the sensitivity of the test, is not conditioned upon the infection status of the tested individual [4,5]. Regions of high disease burden may therefore prioritize test sensitivity, in contrast to a lower burden location’s preference for high test specificity and PPV, all else being equal.

At the heart of an outbreak detection system is a surveillance program that enumerates the baseline rate of case incidence and defines an outbreak as a time period with anomalously high incidence relative to that baseline [9–12]. As many clinical signs and symptoms reflect generic host responses to infection e.g., febrile rash, and infection with a given pathogen can give rise to a wide range of disease symptoms and severity across individuals, accurate methods of case identification are required. Given the imperfect nature of diagnostic classification, any result for an individual is uncertain. Accumulating multiple individual test results to produce population-level counts will propagate this uncertainty, and may result in over- or under-counts due to a preponderance of the diagnostic test to produce either false positive or false negative individual test results. When the prevalence of a surveillance program’s target disease is low relative to the prevalence of other sources of clinically-compatible cases (as might be expected at the start of an outbreak), the PPV of an individual diagnostic will decrease, increasing the number of false positives, making it harder to identify true anomalies in disease incidence. As a result, it has been commonplace for infectious disease surveillance systems to be developed around high-accuracy tests, such as PCR and ELISA tests, when financially and logistically feasible [13–18].

Outbreak detection systems, like diagnostic tests, must prioritize the sensitivity or specificity of an alert to detect an outcome (the outbreak) [19–21]. For many disease systems, particularly in resource constrained environments where the burden of infectious diseases is typically highest [22,23], cases are counted and if a pre-determined threshold is breached — be that weekly, monthly — or some combination of the two, an alert is triggered that may launch a further investigation and/ or a response [20,24]. In effect, this converts a continuous phenomenon (observed cases) into a binary measure (outbreak or no outbreak) for decision making purposes. For reactive responses such as vaccination campaigns and non-pharmaceutical based interventions that are designed to reduce transmission or limit and suppress outbreaks, early action has the potential to avert the most cases [25–30]. While this framing would point towards a sensitive (i.e., early alert) surveillance system being optimal, each action comes with both direct and indirect financial and opportunity costs stemming from unnecessary activities that limit resource availability for future responses. Much like the need to carefully evaluate the balance of an individual diagnostic test’s sensitivity and specificity, it is essential to consider these characteristics at the outbreak level.

The concept of using incidence-based alert triggers to detect the discrete event of an outbreak with characteristics analogous to individual tests has been well documented in the case of meningitis, measles, and malaria [21,24,29,31–34]. However, an overlooked, yet critical, aspect of an outbreak detection system is the interplay between the individual test and outbreak alert characteristics. With their success within malaria surveillance systems, and particularly since the COVID-19 pandemic, rapid diagnostic tests (RDTs) have garnered wider acceptance, and their potential for use in other disease systems has been gaining interest [35]. Despite concerns about their lower diagnostic accuracy slowing their adoption [36], the reduced cold-chain requirements [37], reduced training and laboratory requirements and costs [18,24,37], and more rapid results provided by RDTs relative to ELISAs have been shown to outweigh the cost of false positive/ negative results in some settings [35,38–40].

We examine how the use of imperfect diagnostic tests affects the performance of outbreak detection in the context of measles where RDTs are being developed with promising results [35,37,41,42] (though not exclusively [43]). We evaluate the scenarios under which equivalence in outbreak detection can be achieved, where altering testing rates can offset the reduction in diagnostic discrimination of imperfect tests relative to perfect tests, and meaningful improvements can be attained with respect to specific metrics e.g., speed of response. By examining the combination of the alert threshold and individual test characteristics in a modeling study that explicitly incorporates dynamical background noise, we illustrate the need to develop TPPs for surveillance programs as a whole.

## Results

The threshold that maximized measles surveillance accuracy depends on the diagnostic test characteristics, the testing rate, and the structure of the non-measles noise (Supplemental Table 1, Figure 1). When the average noise incidence was 8 times higher than the average measles incidence (Λ(8)), the optimal outbreak alert threshold (T_O_) ranged between 0.39 and 16.80 test positive cases per day. Not surprisingly, the biggest driver of this difference was the testing rate; as a larger fraction of suspected cases are tested, the optimal threshold generally increases monotonically for all test and noise types with the exception of high dynamical noise scenarios (Supplemental Table 1, Figure 1).

**Figure 1:**
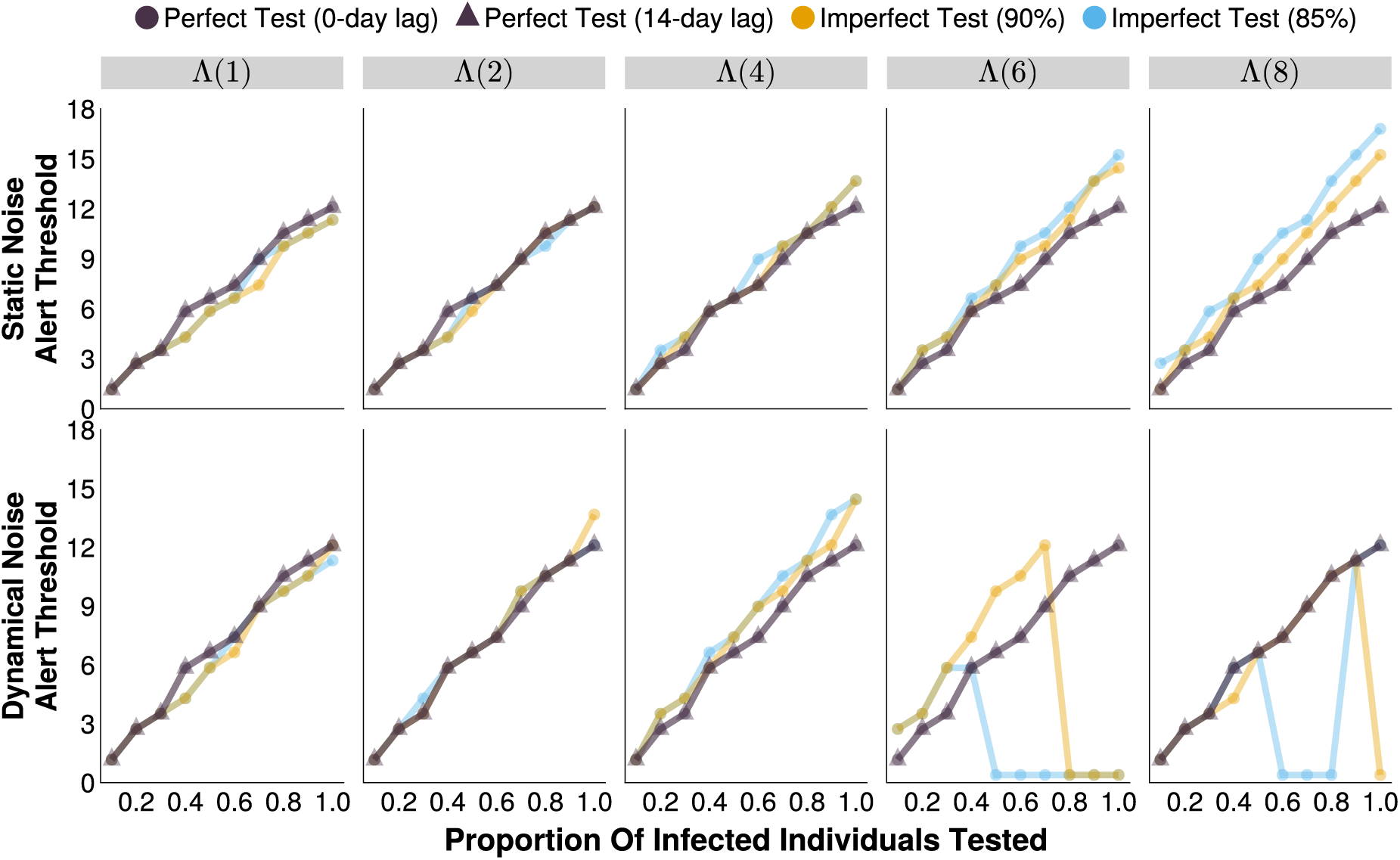
The optimal alert threshold of outbreak detection systems (that maximizes outbreak detection accuracy) under different testing rates and noise structures. Each imperfect test uses the same value for both its sensitivity and specificity (either 85% or 90%). Circular markers represent tests with 0-day turnaround times, and triangular markers represent tests with result delays. Λ(4) indicates the mean noise incidence is 4 times higher than the mean measles incidence, for example. Supplemental Table 1 provides the underlying values in a table format to help distinguish between lines that may overlap.

The maximal attainable surveillance accuracy at the optimal threshold depends strongly on the structure and magnitude of the background noise (Figure 2). For static noise, at all magnitudes, the maximum surveillance accuracy was consistently ≈ 90% accuracy for all diagnostic tests (Figure 2). For dynamical SEIR noise, the perfect tests perform identically to the static noise case at all magnitudes (Figure 2). For imperfect diagnostic tests, which have lower individual sensitivity and specificity, the maximal attainable accuracy is lower than the perfect tests for all testing rates (P) at noise magnitude ≥ Λ(2) (Figure 2). Notably, the surveillance accuracy typically declines with more noise and is not consistently improved with higher testing rates as the signal becomes increasingly dominated by false positive test results (Figure 2). At high levels of dynamical noise (≥ Λ(6)), high testing rates (≥ 50%) result in a marked increase in outbreak detection accuracy. However, in these scenarios the optimal outbreak alert threshold falls to 0.39 (daily) test positive cases in a 7-day moving average: 3 positive test cases in a week would be sufficient to trigger an outbreak alert (Figure 1, Figure 2, Supplemental Table 1).

**Figure 2:**
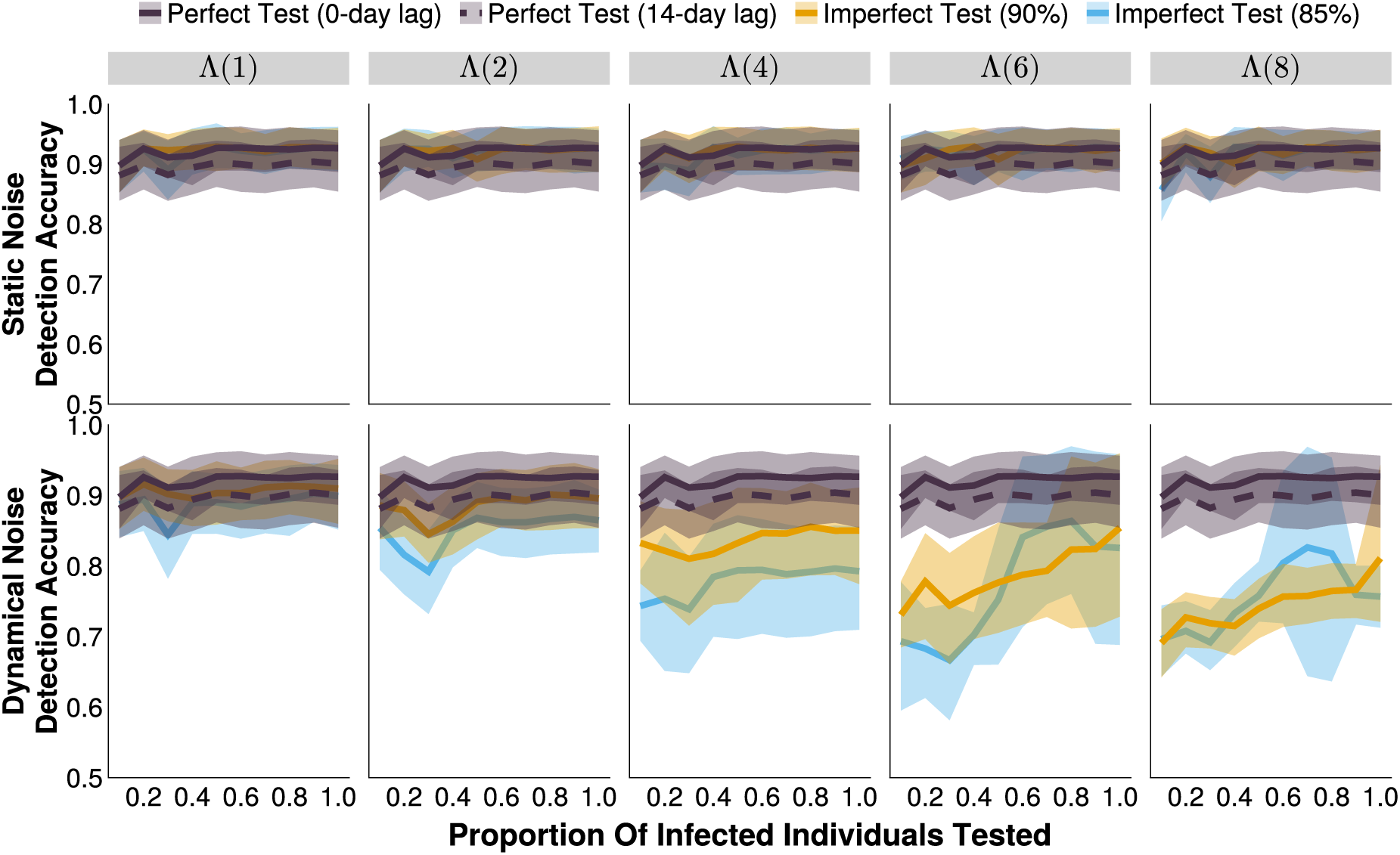
The accuracy of outbreak detection systems under different testing rates and noise structures, at their respective optimal alert thresholds. The shaded bands illustrate the 80% central interval, and the solid/dashed lines represent the mean estimate. Each imperfect test uses the same value for both its sensitivity and specificity (either 85% or 90%). Solid lines represent tests with 0-day turnaround times, and dashed lines represent tests with result delays. Λ(4) indicates the mean noise incidence is 4 times higher than the mean measles incidence, for example.

Introducing a lag in test result reporting can decrease surveillance accuracy. This will occur if an alert occurs within the duration of the lag (e.g., 14 days) of the end of the outbreak. If it is the first alert, then it will be recorded as an outbreak for which there was no alert, reducing the proportion of outbreaks that are correctly identified (our definition of sensitivity). If it is a subsequent alert (recall that multiple alerts may occur within a single outbreak), then it will be recorded as an alert for which there was no outbreak, reducing the proportion of alerts that occur during an outbreak (our definition of positive predictive value). This will disproportionately affect shorter outbreaks. For the conditions simulated here, introducing a 14-day lag in test reporting for a perfect test reduces the surveillance accuracy by ≈ 3% by reducing the PPV, but not the sensitivity, of the system (Supplemental Figure 4, Supplemental Figure 5). In a system with static noise, imperfect tests can achieve slightly higher accuracy than perfect, lagged tests (Figure 2). Given dynamical background noise, perfect, lagged, tests outperform imperfect tests.

A delay in test results reporting does not affect the optimized threshold (T_O_) (Figure 1). However, it always leads to an increase in the median delay from outbreak start to alert, relative to a perfect test with no result delays, as well as imperfect tests (Figure 3). Under high dynamical noise (≥ Λ(6)) and high testing rates (≥ 50%) with imperfect tests, the extremely low optimal threshold results in a very sensitive alert system and large negative delays i.e., the alerts are triggered by false positive test results before the start of the outbreak (Figure 3).

**Figure 3:**
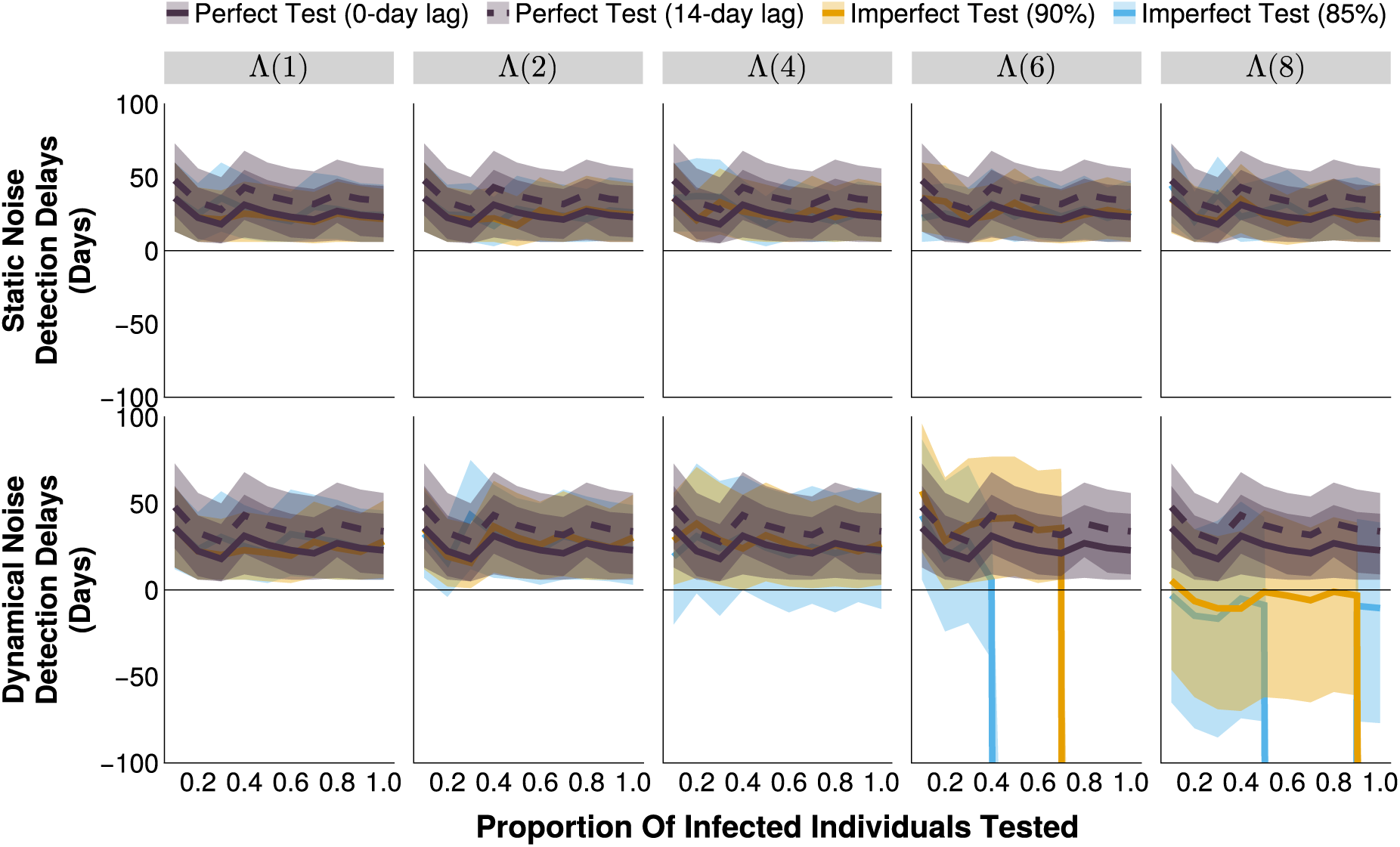
The detection delay of outbreak detection systems under different testing rates and noise structures, at their respective optimal alert thresholds. The shaded bands illustrate the 80% central interval, and the solid/dashed lines represent the mean estimate. Each imperfect test uses the same value for both its sensitivity and specificity (either 85% or 90%). Solid lines represent tests with 0-day turnaround times, and dashed lines represent tests with result delays. Λ(4) indicates the mean noise incidence is 4 times higher than the mean measles incidence.

It is notable that surveillance metrics do not change monotonically with an increase in testing rate, and this holds regardless of the type of test. This effect is more exaggerated for some metrics — detection delays, proportion of time in alert (i.e., the proportion of the simulated time series where the number of test positive cases exceeds the outbreak alert threshold), and number of unavoidable cases (i.e., cases that occur between the outbreak start and its detection) — than for others (accuracy). In general, the increase in accuracy with higher testing rates is accompanied with longer testing delays. This reflects the change from highly sensitive systems with low thresholds to more specific systems with higher thresholds at higher testing rates. For static noise, similar detection delays are observed for all test and noise magnitudes, with most of the variation attributable to the change in the testing rate (means of 24.0 to 36.3 days). Under dynamical noise, a difference in the performance of perfect and imperfect diagnostic tests arise under high magnitudes of dynamical noise (≥ Λ(6)) (Figure 3). As the optimal threshold for imperfect tests drops to ≈ 0, frequent alerts translate to large negative detection delays (≪ −100 days). Negative delays indicate that alerts are being triggered before the start of the outbreak and is correlated with a greater proportion of the time series that is under alert, resulting from fewer, but far longer, alert periods (Figure 4, Supplemental Figure 1, Supplemental Figure 2). Long detection delays can manifest as large numbers of unavoidable cases (Supplemental Figure 3). However, the large negative detection delays associated with high dynamical noise regimes and imperfect diagnostic tests can counter intuitively produce large numbers of unavoidable cases, too, as long, sustained alerts translate to “missed” outbreaks as each alert can only “detect” one outbreak.

**Figure 4:**
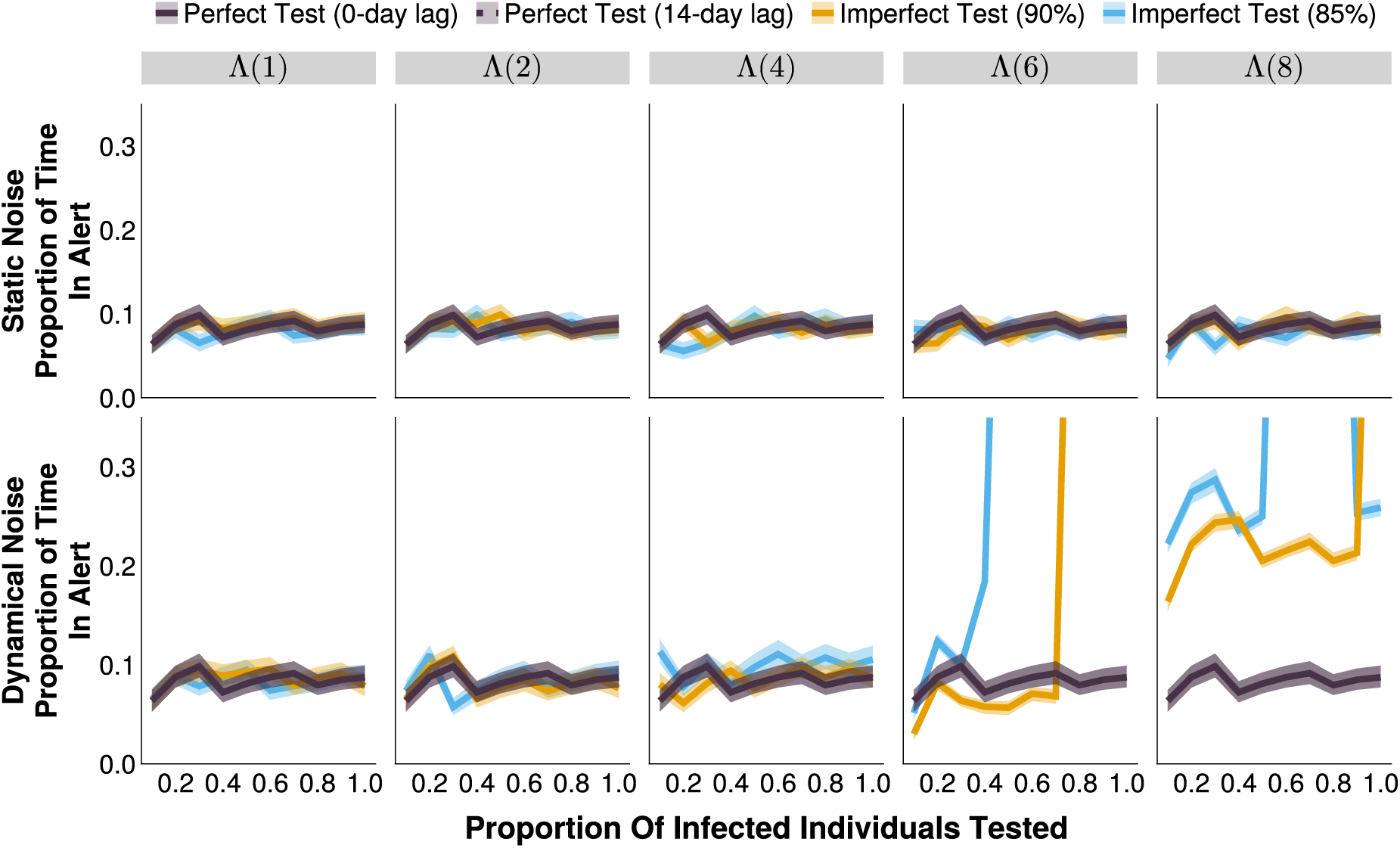
The proportion of the time series in alert of outbreak detection systems under different testing rates and noise structures, at their respective optimal alert thresholds. The shaded bands illustrate the 80% central interval, and the solid/dashed lines represent the mean estimate. Each imperfect test uses the same value for both its sensitivity and specificity (either 85% or 90%). Solid lines represent tests with 0-day turnaround times, and dashed lines represent tests with result delays. Λ(4) indicates the mean noise incidence is 4 times higher than the mean measles incidence.

## Discussion

The performance of an outbreak detection system is highly sensitive to the structure and level of background noise in the simulation. Despite setting the mean daily noise incidence to equivalent values for the dynamical and static simulations, we observed drastically different results.

Under the assumption that non-measles febrile rash is relatively static in time (static noise scenarios), imperfect (RDT-like) diagnostics can perform as well as, if not better than, perfect (ELISA-like) tests, with respect to outbreak detection accuracy, delays, and the number of unavoidable cases, at all testing rates. RDTs for measles are expected to be less expensive than ELISAs, which could lead to overall savings in surveillance systems at a given testing rate, and/or may allow for higher testing rates for the same or lower cost in resource limited settings [37]. However, if it is expected that the noise is dynamic and substantially larger in magnitude than the target infection (≥ Λ(4)), imperfect tests cannot overcome their accuracy limitations through higher testing rates, saturating at c. 80% accuracy, relative to 93% achieved with perfect tests. This discrepancy occurs because, despite the same average incidence of noise in each (comparable) scenario, the relative proportion of measles to noise on any day varies throughout the dynamical noise time series, exacerbating the increase in false positive and negative test results as the diagnostic’s sensitivity and specificity declines. Under extremely high dynamical noise levels (≥ Λ(6)), the relative paucity of true outbreak periods to non-outbreak periods creates a severely imbalanced data set (c. 14% of the time series is within an outbreak period), such that the optimal detection strategy with imperfect diagnostic tests and moderately high testing rates is to maintain a perpetual alert status. In a realistic scenario, this is clearly not a feasible outbreak detection and response strategy.

Surveillance is used to inform action [44]. What actions are taken depend upon the constraints imposed, and the values held, within a particular surveillance context. This analysis is therefore not a complete optimization, which would require explicit decisions to be made about the preference for increased speed at the cost of higher false alert rates and lower PPV (and vice versa). These will be country-specific decisions, and they may change throughout time; for example, favoring RDTs when there are low levels of background infections (either static or dynamical in nature), and ELISAs during large (suspected) non-measles outbreaks of febrile rash illness e.g., rubella. These trade-offs must be explicitly acknowledged when designing surveillance systems, and we present a framework to account for the deep interconnectedness of individual and population-level uncertainties that arise from necessary categorizations.

### Limitations and Strengths

To our knowledge, this is one of the first simulation studies to examine the relationship between individual test characteristics and the wider surveillance program. By explicitly modeling the interaction between the two, we illustrate the dependency of the performance of the surveillance system at the population level and on the characteristics of the diagnostic tests at the individual level. Thus, a change to the latter (e.g., adoption of a new diagnostic with different sensitivity and specificity) without a corresponding change to surveillance frequency or action thresholds, may lead to a reduction in outbreak detection performance. Additionally, by defining outbreak bounds concretely we have been able to calculate metrics of outbreak detection performance that draw parallels to those used when evaluating individual diagnostic tests. This provides an intuitive understanding and simplifies the implementation of this method in resource-constrained environments, something that may not be possible with many outbreak detection and early warning system simulations in the literature. An evaluation of all outbreak detection algorithms is beyond the scope of this work, but a more computationally expensive approach based on nowcasting incidence may help overcome the shortcomings of imperfect diagnostics in high-noise scenarios.

While a simulation-based approach allows for complete determination of true infection status i.e., measles vs non-measles febrile rash cases, and therefore an accurate accounting of the outbreak and alert bounds, these simulations do not specifically represent any real-world setting. The evaluation of empirical data provides this opportunity, but at the cost of not knowing the true infection status of individuals, confounding of multiple variables, limiting analysis to only those who are observed (i.e., not those in the community who do not visit a healthcare center), and removing the possibility to explore the sensitivity of the results when adjusting parameters that are central to a surveillance program e.g., testing rate, and the test itself.

Additionally, it has been well documented that the performance of an individual test is highly sensitive to its timing within a person’s infection cycle [13,18,39,40,45–47], so it is possible that different conclusions would be drawn if temporal information about test administration was included in the simulation. For example, if there is systemic sampling bias such that during the initial exponential phase of the outbreak infectious individuals are ‘captured’ by the surveillance program earlier in their infection cycle (as a growing epidemic has a disproportionately large number of ‘young infections’ [48]), RDTs may have higher accuracy during the earlier phase of the outbreak relative to after the peak. Under these conditions, a diagnostic test that is ‘less accurate’ on average may still have utility for the purposes of detecting an outbreak under high levels of dynamical noise, which only relies on capturing the epidemic’s growth past a given threshold value, in the context of this study. However, this assumes that the number of false positive test results generated decreases due to higher accuracy (including specificity), rather than increasing the diagnostic’s sensitivity being the sole change. Future work should aim to capture these dynamics and characterize the resulting effect on an imperfect diagnostic test’s utility for outbreak detection in a regime with high dynamical noise.

Finally, despite numerous scenarios where equivalent outbreak detection accuracy could be achieved, under regimes with high levels of dynamical noise, imperfect tests were not able to appropriately distinguish between outbreak and non-outbreak periods. In these situations, the added complexity from large numbers of false positive test results likely warrants a different decision criteria than a binary detection threshold. Similarly, the optimal threshold depends heavily on the costs ascribed to incorrect actions, be that failing to detect an outbreak or incorrectly mounting a response for an outbreak that does not exist. In the simulations we have weighted them equally (as the system’s accuracy is defined as the arithmetic mean of the sensitivity and PPV), but it is likely that they should not be deemed equivalent; missing an outbreak may result in many thousands of cases, whereas an unnecessary alert would generally launch an initial low-cost investigation for full determination of the outbreak status. This is particularly important in countries with vast heterogeneity in transmission: different weightings should be applied to higher vs. lower priority/ risk regions to account for discrepancies in the consequences of incorrect decisions.

Given these limitations, the explicit values (i.e., optimal thresholds, accuracies etc.) should be interpreted with caution, and the exact results observed in the real-world will likely be highly dependent on unseen factors, such as the proportion of measles and non-measles febrile rash patients that seek healthcare. However, the general pattern that imperfect tests can produce equivalent outbreak detection capabilities under static or low dynamical noise regimes, should hold. More importantly, the analysis framework provides a consistent and holistic approach to evaluating the trade-off between individual level tests and the alert system enacted to detect outbreaks.

## Methods

### Model Structure

We constructed a stochastic compartmental non-age structured Susceptible-Exposed-Infected-Recovered (SEIR) model of measles, and simulated disease transmission using a modified Tau-leaping algorithm with a time step of 1 day [49]. We utilized binomial draws to ensure compartment sizes remained positive valued [50]. We assumed that the transmission rate (*β*_*t*_) is sinusoidal with a period of one year and 20% seasonal amplitude. *R*_0_ was set to 16, with a latent period of 10 days and infectious period of 8 days [13,51]. The population was initialized with 500,000 individuals with Ghana-like birth and vaccination rates, and the final results were scaled up to the approximate 2022 population size of Ghana (33 million) [52]. Ghana was chosen to reflect a setting with a high-performing measles vaccination program that has not yet achieved elimination status (c. 80% coverage for two doses of measles-containing vaccine), and must remain vigilant to outbreaks [53,54]. We assumed commuter-style imports at each time step to avoid extinction; the number of imports each day were drawn from a Poisson distribution with mean proportional to the size of the population and *R*_0_ [55]. The full table of parameters can be found in Table 1. All simulations and analyses were completed in Julia version 1.11.5 [56], with all code stored at https://github.com/ arnold-c/OutbreakDetection.

**Table 1:**
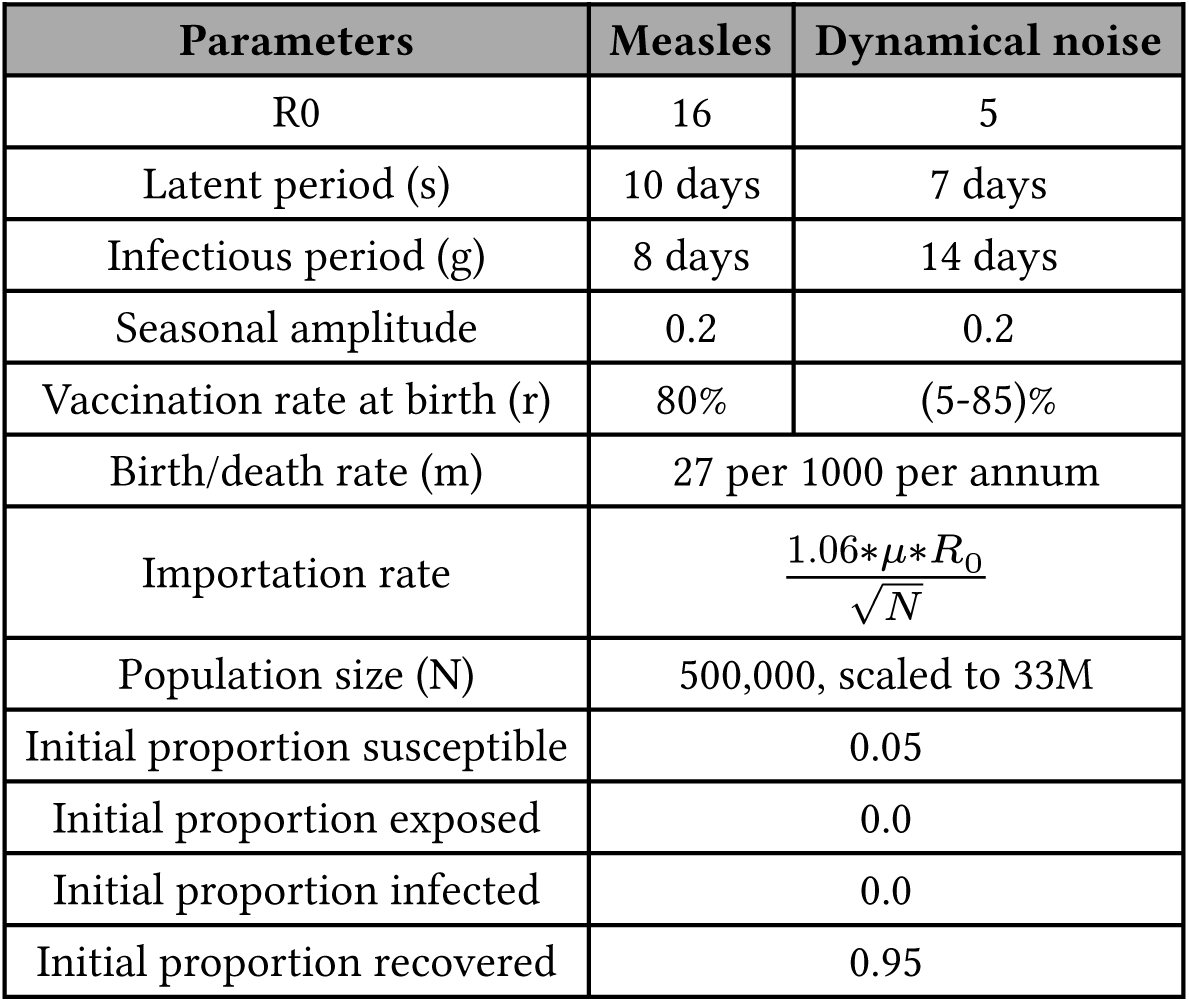
Compartmental model parameters.

| Parameters | Measles | Dynamical noise |
| --- | --- | --- |
| $R_0$ | 16 | 5 |
| Latent period (s) | 10 days | 7 days |
| Infectious period (g) | 8 days | 14 days |
| Seasonal amplitude | 0.2 | 0.2 |
| Vaccination rate at birth (r) | 80% | (5-85)% |
| Birth/death rate (m) | 27 per 1000 per annum |  |
| Importation rate | $\frac{1.06 * \mu * R_0}{\sqrt{N}}$ | |
| Population size (N) | 500,000, scaled to 33M |  |
| Initial proportion susceptible | 0.05 |  |
| Initial proportion exposed | 0.0 |  |
| Initial proportion infected | 0.0 |  |
| Initial proportion recovered | 0.95 |  |

To examine the sensitivity of the detection system to background noise, we generated a time series of symptomatic febrile rash by combining the measles incidence time series with a noise time series. The noise time series was modeled as either Poisson-only noise (subsequently referred to as static noise), to represent the incidence of non-specific febrile rash due to any of a number of possible etiologies, or dynamical noise modeled as a rubella SEIR process. For static noise, the time series of non-measles febrile rash cases each day was constructed by independent draws from a Poisson distribution. For dynamical noise, we generated time series of cases from an SEIR model that matched the measles model in structure, but had *R*_0_ = 5, mean latent period of 7 days, and mean infectious period of 14 days. We also added additional static noise drawn from a Poisson distribution with mean equal to 15% of the average daily rubella incidence to account for non-rubella sources of febrile rash (Table 1) [57,58]. The seasonality for the rubella noise was simulated to be in-phase with measles.

For each noise structure, we simulated five magnitudes of noise (Λ), representing the average daily noise incidence. Λ was calculated as a multiple (*c*) of the average daily measles incidence (⟨Δ*I*_*M*_ ⟩): Λ = *c* ⋅ ⟨Δ*I*_*M*_ ⟩ where *c* ∈ {1, 2, 4, 6, 8}. Noise magnitudes will be denoted as Λ(*c*) for the rest of the manuscript e.g., Λ(8) to denote scenarios where the average noise incidence is 8 times that of the average measles incidence. For the static noise scenarios, independent draws from a Poisson distribution with mean *c* ⋅ ⟨Δ*I*_*M*_ ⟩ were simulated to produce the noise time series i.e., Λ(*c*) = Pois(*c* ⋅ ⟨Δ*I*_*M*_ ⟩). For the dynamical noise scenarios, the rubella vaccination rate at birth was set to 85.38%, 73.83%, 50.88%, 27.89%, or 4.92% to produce equivalent values of Λ (to within 2 decimal places): Λ(*c*) = ⟨Δ*I*_*R*_⟩ + Pois(0.15 ⋅ ⟨Δ*I*_*R*_⟩). We simulated 100 time series of 100 years for each scenario before summarizing the distributions of outbreak detection methods.

### Defining Outbreaks

It is common to use expert review to define outbreaks when examining empirical data, but this is not feasible in a modeling study where tens of thousands of years are being simulated. Previous simulation studies define an outbreak as a period where *R*_t_ > 1 with the aim of detecting an outbreak during the grow period [59,60], or use a threshold of > 2 standard deviations (s.d.) over the mean seasonal incidence observed in empirical data (or from a ‘burn-in’ period of the simulation) [61–64].

Here we simulate time series of 100 years and we define a measles outbreak as a region of the time series that meets the following three criteria:

- The daily measles incidence must be greater than or equal to 5 cases
- The daily measles incidence must remain above 5 cases for greater than or equal to 30 consecutive days
- The total measles incidence must be great than or equal to 500 cases within the bounds of the outbreak

Only events meeting all 3 criteria are classified as outbreaks. The incidence of non-measles febrile rash (i.e., noise) does not affect the outbreak status of a region but may affect the alert status triggered by the testing protocol.

Each day, a percentage (P) of clinically-compatible cases of febrile rash are tested; P is fixed in a given scenario to a value between 10% and 100%, in 10% increments. Each “testing scenario” combines a testing rate (P) with one of the following tests:

- An imperfect test with 85% sensitivity and specificity, and 0-day lag in result return. That is, 85% of true measles cases will be correctly labeled as positive, and 15% of non-measles febrile rash individuals that are tested will be incorrectly labeled as positive for measles. This acts as a lower bound of acceptability for a hypothetical measles RDT [41]
- An imperfect test with 90% sensitivity and specificity, and 0-day lag in result return [37]
- A perfect test with 100% sensitivity and specificity, and a 0-day test result delay. This is more accurate than is observed for current ELISA tests [65], but it used to evaluate the theoretical best-case scenario
- A perfect test with 100% sensitivity and specificity, and a 14-day test result delay that represents a best-case test under more realistic reporting delays in result return

For each time series of true measles cases, we define outbreaks as the range of time that meets the definition above (Figure 5a). We then add non-measles noise (Figure 5b) and test according to the testing scenario, which yields 5 time series of test positive cases (Figure 5c): one time series of all clinically compatible cases and 4 reflecting the testing scenarios.

**Figure 5:**
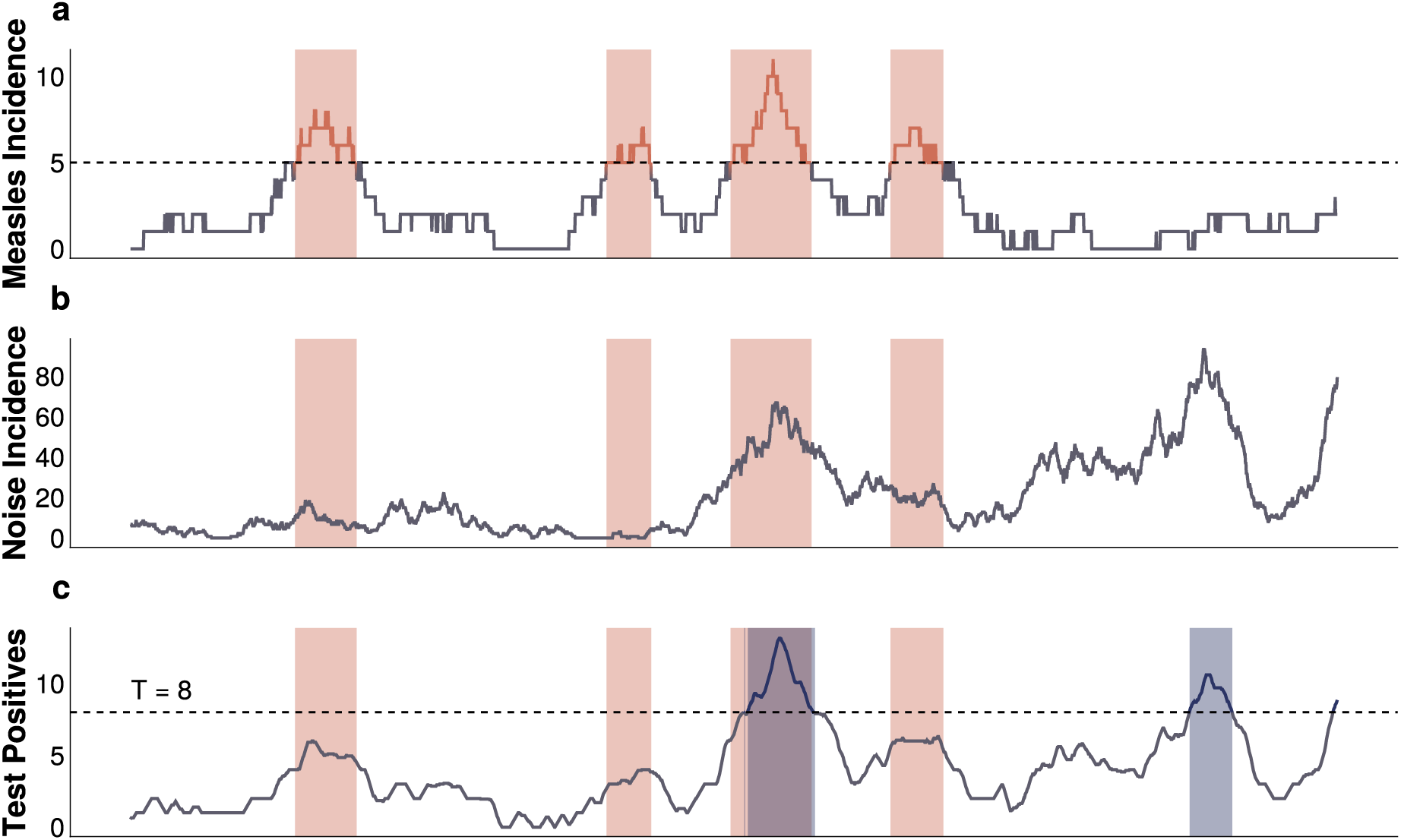
A schematic of the outbreak definition and alert detection system. A) Measles incidence time series. B) Noise incidence time series. C) Observed time series of test positive cases according to a given testing scenario. The orange bands present in all 3 panels represent regions of the measles time series that meet the outbreak definition criteria. In panel C, the dark blue bands represent regions of the test positive time series that breach the alert threshold (the horizontal dashed line), and constitute an alert.

### Triggering Alerts

We define an “alert” as any consecutive string of 1 or more days where the 7-day (trailing) moving average of the test positive cases is greater than or equal to a pre-specified alert threshold, T. For each time series of test positive cases, we calculate the percentage of alerts that are “correct”, defined as any overlap of 1 or more days between the outbreak and alert periods (Figure 5c). This is analogous to the PPV of the alert system and will be referred to as such for the rest of the manuscript. It is possible to have multiple alerts within a single outbreak if the 7-day moving average of test positive cases drops below, and then re-crosses, the threshold, T, and we count each as correct. For all outbreaks in the measles time series, we calculate the percentage that contain at least 1 alert within the outbreak’s start and end dates (Figure 5c). We refer to this as the sensitivity of the alert system. We also calculate the detection delay as the time from the start of an outbreak to the start of its first alert. If the alert period starts before the outbreak and continues past the start date of the outbreak, this would be considered a correct alert with a negative delay i.e., an early warning triggered by false positive test results. Finally, for each time series we calculate the number of unavoidable and avoidable outbreak cases. Unavoidable cases are those that occur before a correct alert, or those that occur in an undetected outbreak. Avoidable cases are those that occur within an outbreak and after the first alert; we do not quantity avoidable cases here as the value depends critically on the explicit details of the response, which we do not model.

We define the accuracy of the surveillance system for a given time series as the arithmetic mean of the system’s PPV and sensitivity. To examine the interaction of the test with the surveillance system’s characteristics (i.e., testing rate, noise structure and magnitude), we optimized the alert threshold, T, for a given “testing scenario”. Each of the 100 simulations per scenario produces an accuracy, and we identified the optimal alert threshold, T_O_, as the value that produced the highest mean accuracy for a given scenario. To identify T_O_ we implemented the TikTak multistart optimization algorithm [66], using 100 initial values (alert thresholds) selected from a Sobol’ low-discrepancy sequence [67] initialized with lower and upper bounds of 0.0 and 50.0, respectively. In brief, the Sobol’ sequence is a deterministic, quasi-random sequence of numbers that maximizes the uniformity of the explored parameter space by approximately iteratively bisecting the parameter space [67,68]. After 100 initial alert thresholds are generated, the accuracy is evaluated and the 10 alert thresholds (points) with the highest accuracy are retained. The 10 retained alert thresholds are sorted in descending order of accuracy, creating the sequence of Sobol’ points (**s_1_…s_10_**) that are used to calculate the seed points for local optimization that subsequently performed using the BOBYQA derivative-free algorithm [69]. For each of the 10 local optimizations, the starting seed is computed as the weighted combination of the Sobol’ point **s_i_** and the alert threshold that produced the maximum accuracy so far, with increasing weight provided to the alert threshold that maximized accuracy; more information can be found in Appendix B.6 of [66]. The TikTak algorithm is implemented in the MultistartOptimization.jl package [70], with local optimization (BOBYQA) implemented in the NLOpt.jl package [71].

We then compare testing scenarios at their respective optimal alert threshold. This allows for conclusions to be made about the surveillance system as a whole, rather than just single components. We also present results for optimizations based upon the harmonic mean (F-1 score) of the system’s PPV and sensitivity in the Supplement (Supplemental Figure 6, Supplemental Figure 7, Supplemental Figure 8, Supplemental Figure 9).

## Author Contributions

*Conceptualization:* all authors

*Data curation:* CA, MJF

*Formal analysis:* CA

*Funding acquisition:* ACK, AW, BP, MJF, WJM

*Investigation:* CA

*Methodology:* CA, MJF

*Project administration:* MJF

*Software:* CA

*Supervision:* AW, BP, MJF, WJM

*Validation:* CA

*Visualization:* CA

*Writing - original draft:* CA

*Writing - review and editing:* all authors

## Data Access, Responsibility, and Analysis

Callum Arnold and Dr. Matthew J. Ferrari had full access to all the data in the study and take responsibility for the integrity of the data and the accuracy of the data analysis. Callum Arnold (Department of Biology, Pennsylvania State University) conducted the data analysis.

## Data Availability

All code and data for the simulations can be found at https://github.com/arnold-c/OutbreakDetection

## Bibliography

[1] World Health Organization. Diagnostics 2024.

[2] Yang S, Rothman RE. PCR-based Diagnostics for Infectious Diseases: Uses, Limitations, and Future Applications in Acute-Care Settings. The Lancet Infectious Diseases 2004;4:337. 10.1016/S1473-3099(04)01044-8.

[3] Alhajj M, Zubair M, Farhana A. Enzyme Linked Immunosorbent Assay. StatPearls 2024.

[4] Westreich D. Diagnostic Testing, Screening, and Surveillance. Epidemiology by Design: A Causal Approach to the Health Sciences 2019:0. 10.1093/oso/9780190665760.003.0005.

[5] Shreffler J, Huecker MR. Diagnostic Testing Accuracy: Sensitivity, Specificity, Predictive Values and Likelihood Ratios. StatPearls 2024.

[6] Parikh R, Mathai A, Parikh S, Chandra Sekhar G, Thomas R. Understanding and Using Sensitivity, Specificity and Predictive Values. Indian Journal of Ophthalmology 2008;56:45–50.

[7] World Health Organization. Target Product Profiles 2024.

[8] Chua AC, Cunningham J, Moussy F, Perkins MD, Formenty P. The Case for Improved Diagnostic Tools to Control Ebola Virus Disease in West Africa and How to Get There. PLOS Neglected Tropical Diseases 2015;9:e3734. 10.1371/journal.pntd.0003734.

[9] Murray J, Cohen AL. Infectious Disease Surveillance. International Encyclopedia of Public Health 2017:222–9. 10.1016/B978-0-12-803678-5.00517-8.

[10] Zhou X-N, Bergquist R, Tanner M. Elimination of Tropical Disease through Surveillance and Response. Infectious Diseases of Poverty 2013;2:1. 10.1186/2049-9957-2-1.

[11] PAHO. An Integrated Approach to Communicable Disease Surveillance. 2000.

[12] Cragg L. Outbreak Response. Applied Communicable Disease Control 2018:134–51.

[13] Gastanaduy PA, Redd SB, Clemmons NS, Lee AD, Hickman CJ, Rota PA, et al. Measles. Manual for the Surveillance of Vaccine-Preventable Diseases 2019.

[14] Commissioner O of the. Coronavirus (COVID-19) Update: FDA Informs Public About Possible Accuracy Concerns with Abbott ID NOW Point-of-Care Test 2020.

[15] Grassly NC, Pons-Salort M, Parker EPK, White PJ, Ferguson NM, Imperial College COVID-19 Response Team. Comparison of Molecular Testing Strategies for COVID-19 Control: A Mathematical Modelling Study. The Lancet Infectious Diseases 2020;20:1381–9. 10.1016/S1473-3099(20)30630-7.

[16] Ezhilan M, Suresh I, Nesakumar N. SARS-CoV, MERS-CoV and SARS-CoV-2: A Diagnostic Challenge. Measurement 2021;168:108335. 10.1016/j.measurement.2020.108335.

[17] World Health Organization. Cholera 2023.

[18] Essential Programme on Immunization (EPI), Immunization, Vaccines and Biologicals (IVB). Clinical Specimens for the Laboratory Confirmation and Molecular Epidemiology of Measles, Rubella, and CRS. Manual for the Laboratory-based Surveillance of Measles, Rubella, And Congenital Rubella Syndrome 2018.

[19] German RR. Sensitivity and Predictive Value Positive Measurements for Public Health Surveillance Systems. Epidemiology 2000;11:720–7.

[20] World Health Organization. Operational Thresholds. Meningitis Outbreak Response in Sub-Saharan Africa: WHO Guideline 2014.

[21] Lewis R, Nathan N, Diarra L, Belanger F, Paquet C. Timely Detection of Meningococcal Meningitis Epidemics in Africa. The Lancet 2001;358:287–93. 10.1016/S0140-6736(01)05484-8.

[22] GBD 2019 Child and Adolescent Communicable Disease Collaborators. The Unfinished Agenda of Communicable Diseases among Children and Adolescents before the COVID-19 Pandemic, 1990–2019: A Systematic Analysis of the Global Burden of Disease Study 2019. The Lancet 2023;402:313–35. 10.1016/S0140-6736(23)00860-7.

[23] Roser M, Ritchie H, Spooner F. Burden of Disease. Our World in Data 2023.

[24] World Health Organization. Measles Outbreak Guide. Geneva, Switzerland: World Health Organization; 2022.

[25] Atkins BD, Jewell CP, Runge MC, Ferrari MJ, Shea K, Probert WJM, et al. Anticipating Future Learning Affects Current Control Decisions: A Comparison between Passive and Active Adaptive Management in an Epidemiological Setting. Journal of Theoretical Biology 2020;506:110380. 10.1016/j.jtbi.2020.110380.

[26] Tao Y, Shea K, Ferrari M. Logistical Constraints Lead to an Intermediate Optimum in Outbreak Response Vaccination. PLOS Computational Biology 2018;14:20. 10.1371/journal.pcbi.1006161.

[27] Grais R, Conlan A, Ferrari M, Djibo A, Le Menach A, Bjørnstad O, et al. Time Is of the Essence: Exploring a Measles Outbreak Response Vaccination in Niamey, Niger. Journal of the Royal Society Interface 2008;5:67–74. 10.1098/rsif.2007.1038.

[28] Ferrari MJ, Fermon F, Nackers F, Llosa A, Magone C, Grais RF. Time Is (Still) of the Essence: Quantifying the Impact of Emergency Meningitis Vaccination Response in Katsina State, Nigeria. International Health 2014;6:282–90. 10.1093/inthealth/ihu062.

[29] World Health Organization. Confirming, Investigating and Managing an Outbreak. Response to Measles Outbreaks in Measles Mortality Reduction Settings: Immunization, Vaccines and Biologicals 2009;3.

[30] Minetti A, Kagoli M, Katsulukuta A, Huerga H, Featherstone A, Chiotcha H, et al. Lessons and Challenges for Measles Control from Unexpected Large Outbreak, Malawi. Emerging Infectious Diseases 2013;19:202–9. 10.3201/eid1902.120301.

[31] Trotter CL, Cibrelus L, Fernandez K, Lingani C, Ronveaux O, Stuart JM. Response Thresholds for Epidemic Meningitis in Sub-Saharan Africa Following the Introduction of MenAfriVac®. Vaccine 2015;33:6212–7. 10.1016/j.vaccine.2015.09.107.

[32] Cooper LV, Stuart JM, Okot C, Asiedu-Bekoe F, Afreh OK, Fernandez K, et al. Reactive Vaccination as a Control Strategy for Pneumococcal Meningitis Outbreaks in the African Meningitis Belt: Analysis of Outbreak Data from Ghana. Vaccine 2019;37:5657–63. 10.1016/j.vaccine.2017.12.069.

[33] Zalwango MG, Zalwango JF, Kadobera D, Bulage L, Nanziri C, Migisha R, et al. Evaluation of Malaria Outbreak Detection Methods, Uganda, 2022. Malaria Journal 2024;23:18. 10.1186/s12936-024-04838-w.

[34] Kaninda A-V, Belanger F, Lewis R, Batchassi E, Aplogan A, Yakoua Y, et al. Effectiveness of Incidence Thresholds for Detection and Control of Meningococcal Meningitis Epidemics in Northern Togo. International Journal of Epidemiology 2000;29:933–40. 10.1093/ije/29.5.933.

[35] Warrener L, Andrews N, Koroma H, Alessandrini I, Haque M, Garcia CC, et al. Evaluation of a Rapid Diagnostic Test for Measles IgM Detection; Accuracy and the Reliability of Visual Reading Using Sera from the Measles Surveillance Programme in Brazil, 2015. Epidemiology & Infection 2023;151:e151. 10.1017/S0950268823000845.

[36] Miller E, Sikes HD. Addressing Barriers to the Development and Adoption of Rapid Diagnostic Tests in Global Health. Nanobiomedicine 2015;2:6. 10.5772/61114.

[37] Brown DW, Warrener L, Scobie HM, Donadel M, Waku-Kouomou D, Mulders MN, et al. Rapid Diagnostic Tests to Address Challenges for Global Measles Surveillance. Current Opinion in Virology 2020;41:77–84. 10.1016/j.coviro.2020.05.007.

[38] McMorrow ML, Aidoo M, Kachur SP. Malaria Rapid Diagnostic Tests in Elimination Settings— Can They Find the Last Parasite?. Clinical Microbiology and Infection : the Official Publication of the European Society of Clinical Microbiology and Infectious Diseases 2011;17:1624–31. 10.1111/j.1469-0691.2011.03639.x.

[39] Larremore DB, Wilder B, Lester E, Shehata S, Burke JM, Hay JA, et al. Test Sensitivity Is Secondary to Frequency and Turnaround Time for COVID-19 Screening. Science Advances 2021;7:eabd5393. 10.1126/sciadv.abd5393.

[40] Middleton C, Larremore DB. Modeling the Transmission Mitigation Impact of Testing for Infectious Diseases. Science Advances 2024;10:eadk5108. 10.1126/sciadv.adk5108.

[41] FIND. Target Product Profile for Surveillance Tests for Measles and Rubella. Geneva, Switzerland: 2024.

[42] Shonhai A, Warrener L, Mangwanya D, Slibinskas R, Brown K, Brown D, et al. Investigation of a Measles Outbreak in Zimbabwe, 2010: Potential of a Point of Care Test to Replace Laboratory Confirmation of Suspected Cases. Epidemiology and Infection 2015;143:3442–50. 10.1017/S0950268815000540.

[43] Senin A, Noordin NM, Sani JAM, Mahat D, Donadel M, Scobie HM, et al. A Measles IgM Rapid Diagnostic Test to Address Challenges with National Measles Surveillance and Response in Malaysia. PLOS ONE 2024;19:e298730. 10.1371/journal.pone.0298730.

[44] World Health Organization. Surveillance in Emergencies 2024.

[45] Helfand RF, Heath JL, Anderson LJ, Maes EF, Guris D, Bellini WJ. Diagnosis of Measles with an IgM Capture EIA: The Optimal Timing of Specimen Collection after Rash Onset. The Journal of Infectious Diseases 1997;175:195–9.

[46] Kissler SM, Fauver JR, Mack C, Olesen SW, Tai C, Shiue KY, et al. Viral Dynamics of Acute SARS-CoV-2 Infection and Applications to Diagnostic and Public Health Strategies. PLOS Biology 2021;19:e3001333. 10.1371/journal.pbio.3001333.

[47] Ratnam S, Tipples G, Head C, Fauvel M, Fearon M, Ward BJ. Performance of Indirect Immunoglobulin M (IgM) Serology Tests and IgM Capture Assays for Laboratory Diagnosis of Measles. Journal of Clinical Microbiology 2000;38:99–104.

[48] Hay JA, Kennedy-Shaffer L, Kanjilal S, Lennon NJ, Gabriel SB, Lipsitch M, et al. Estimating Epidemiologic Dynamics from Cross-Sectional Viral Load Distributions. Science 2021;373:eabh635. 10.1126/science.abh0635.

[49] Gillespie DT. Approximate Accelerated Stochastic Simulation of Chemically Reacting Systems. The Journal of Chemical Physics 2001;115:1716–33. 10.1063/1.1378322.

[50] Chatterjee A, Vlachos DG, Katsoulakis MA. Binomial Distribution Based Tau-Leap Accelerated Stochastic Simulation. The Journal of Chemical Physics 2005;122:24112. 10.1063/1.1833357.

[51] Guerra FM, Bolotin S, Lim G, Heffernan J, Deeks SL, Li Y, et al. The Basic Reproduction Number (R0) of Measles: A Systematic Review. The Lancet Infectious Diseases 2017;17:e420–8. 10.1016/S1473-3099(17)30307-9.

[52] World Bank. Ghana 2024.

[53] World Health Organization. Measles Vaccination Coverage 2024.

[54] Masresha BG, Wiysonge CS, Katsande R, O’Connor PM, Lebo E, Perry RT. Tracking Measles and Rubella Elimination Progress—World Health Organization African Region, 2022–2023. Vaccines 2024;12:949. 10.3390/vaccines12080949.

[55] Keeling MJ, Rohani P. Modeling Infectious Diseases in Humans and Animals. Princeton: Princeton University Press; 2008.

[56] Bezanson J, Edelman A, Karpinski S, Shah VB. Julia: A Fresh Approach to Numerical Computing. SIAM Review 2017;59:65–98. 10.1137/141000671.

[57] Papadopoulos T, Vynnycky E. Estimates of the Basic Reproduction Number for Rubella Using Seroprevalence Data and Indicator-Based Approaches. Plos Computational Biology 2022;18:e1008858. 10.1371/journal.pcbi.1008858.

[58] Morales M, Lanzieri T, Reef S. Rubella. CDC Yellow Book 2024: Health Information for International Travel 2023.

[59] Jombart T, Ghozzi S, Schumacher D, Taylor TJ, Leclerc QJ, Jit M, et al. Real-Time Monitoring of COVID-19 Dynamics Using Automated Trend Fitting and Anomaly Detection. Philosophical Transactions of the Royal Society B: Biological Sciences 2021;376:20200266. 10.1098/rstb.2020.0266.

[60] Stolerman LM, Clemente L, Poirier C, Parag KV, Majumder A, Masyn S, et al. Using Digital Traces to Build Prospective and Real-Time County-Level Early Warning Systems to Anticipate COVID-19 Outbreaks in the United States. Science Advances 2023;9:eabq199. 10.1126/sciadv.abq0199.

[61] Stern L, Lightfoot D. Automated Outbreak Detection: A Quantitative Retrospective Analysis. Epidemiology and Infection 1999;122:103–10.

[62] Salmon M, Schumacher D, Höhle M. Monitoring Count Time Series in R: Aberration Detection in Public Health Surveillance. Journal of Statistical Software 2016;70. 10.18637/jss.v070.i10.

[63] Teklehaimanot HD, Schwartz J, Teklehaimanot A, Lipsitch M. Alert Threshold Algorithms and Malaria Epidemic Detection. Emerging Infectious Diseases 2004;10:1220–6. 10.3201/eid1007.030722.

[64] Leclère B, Buckeridge DL, Boëlle P-Y, Astagneau P, Lepelletier D. Automated Detection of Hospital Outbreaks: A Systematic Review of Methods. PLOS ONE 2017;12:e176438. 10.1371/journal.pone.0176438.

[65] Hiebert J, Zubach V, Charlton CL, Fenton J, Tipples GA, Fonseca K, et al. Evaluation of Diagnostic Accuracy of Eight Commercial Assays for the Detection of Measles Virus-Specific IgM Antibodies. Journal of Clinical Microbiology 2021;59:e3161. 10.1128/JCM.03161-20.

[66] Arnoud A, Kleineberg T, Guvenen F. Benchmarking Global Optimizers 2023. 10.2139/ssrn.4642330.

[67] Sobol’ IM. On the Distribution of Points in a Cube and the Approximate Evaluation of Integrals. USSR Computational Mathematics and Mathematical Physics 1967;7:86–112. 10.1016/0041-5553(67)90144-9.

[68] Lemieux C. Quasi–Monte Carlo Constructions. Monte Carlo and Quasi-Monte Carlo Sampling 2009:139–200. 10.1007/978-0-387-78165-5.

[69] Powell MJD. The BOBYQA Algorithm for Bound Constrained Optimization without Derivatives. Cambridge, UK: 2009.

[70] Papp TK. MultistartOptimization.Jl 2022.

[71] Johnson SJ. The NLopt Nonlinear-Optimization Package 2025.

