## Supplemental Appendix for "Individual and Population Level Uncertainty Interact to Determine the Performance of Outbreak Surveillance Systems"

#### Tables

Supplemental Table 1: The optimal outbreak alert thresholds for imperfect and perfect diagnostic tests (that maximizes outbreak detection accuracy) under dynamical and static noise structures where the average daily noise incidence is 8 times the average daily measles incidence $\Lambda(8)$. Each imperfect test uses the same value for both its sensitivity and specificity (either 85% or 90%).

| | **Test Characteristic** | | **Testing Rate** | | | | | | | | | |
|---|---|---|---|---|---|---|---|---|---|---|---|---|
| Noise Type | Test Type | Test Lag | 10% | 20% | 30% | 40% | 50% | 60% | 70% | 80% | 90% | 100% |
| Static noise | Imperfect Test (85%) | 0 | 2.73 | 3.52 | 5.86 | 6.64 | 8.98 | 10.55 | 11.33 | 13.67 | 15.23 | 16.8 |
| Static noise | Imperfect Test (90%) | 0 | 1.17 | 3.52 | 4.3 | 6.64 | 7.42 | 8.98 | 10.55 | 12.11 | 13.67 | 15.23 |
| Dynamical noise | Imperfect Test (85%) | 0 | 1.17 | 2.73 | 3.52 | 5.86 | 6.64 | 0.39 | 0.39 | 0.39 | 11.33 | 12.11 |
| Dynamical noise | Imperfect Test (90%) | 0 | 1.17 | 2.73 | 3.52 | 4.3 | 6.64 | 7.42 | 8.98 | 10.55 | 11.33 | 0.39 |
| All noise structures | Perfect Test | 14 | 1.17 | 2.73 | 3.52 | 5.86 | 6.64 | 7.42 | 8.98 | 10.55 | 11.33 | 12.11 |
| All noise structures | Perfect Test | 0 | 1.17 | 2.73 | 3.52 | 5.86 | 6.64 | 7.42 | 8.98 | 10.55 | 11.33 | 12.11 |

### Figures

#### Arithmetic Mean Optimizations

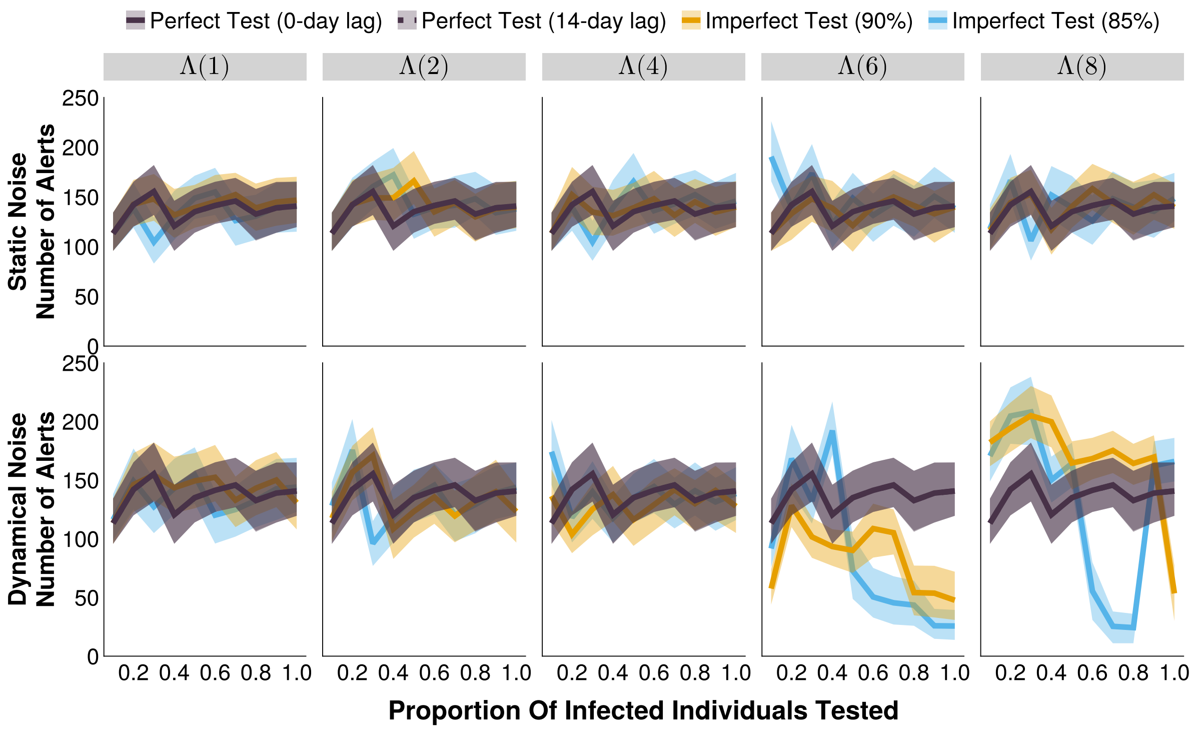


Supplemental Figure 1: The number of alerts of outbreak detection systems under different testing rates and noise structures, at their respective optimal alert thresholds. The shaded bands illustrate the 80% central interval, and the solid/dashed lines represent the mean estimate. Each imperfect test uses the same value for both its sensitivity and specificity (either 85% or 90%). Solid lines represent tests with 0-day turnaround times, and dashed lines represent tests with result delays. $\Lambda(4)$ indicates the mean noise incidence is 4 times higher than the mean measles incidence, for example.

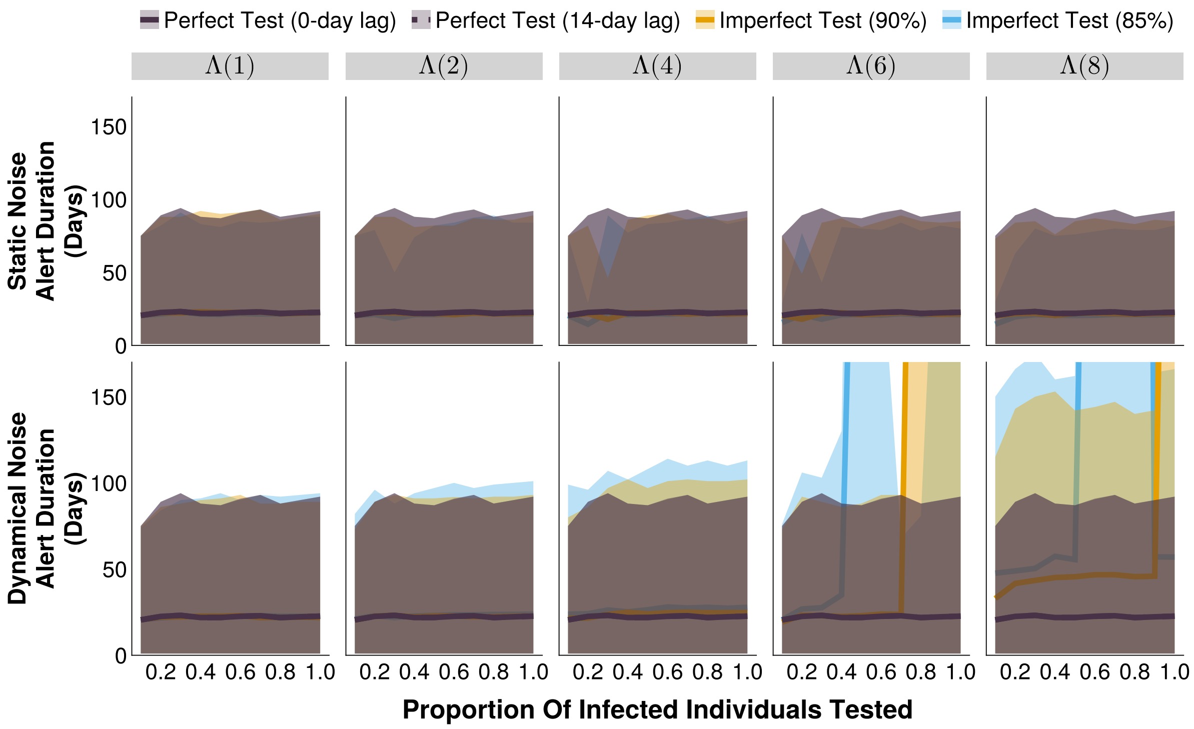


Supplemental Figure 2: The duration of alerts of outbreak detection systems under different testing rates and noise structures, at their respective optimal alert thresholds. The shaded bands illustrate the 80% central interval, and the solid/dashed lines represent the mean estimate. Each imperfect test uses the same value for both its sensitivity and specificity (either 85% or 90%). Solid lines represent tests with 0-day turnaround times, and dashed lines represent tests with result delays. $\Lambda(4)$ indicates the mean noise incidence is 4 times higher than the mean measles incidence, for example.

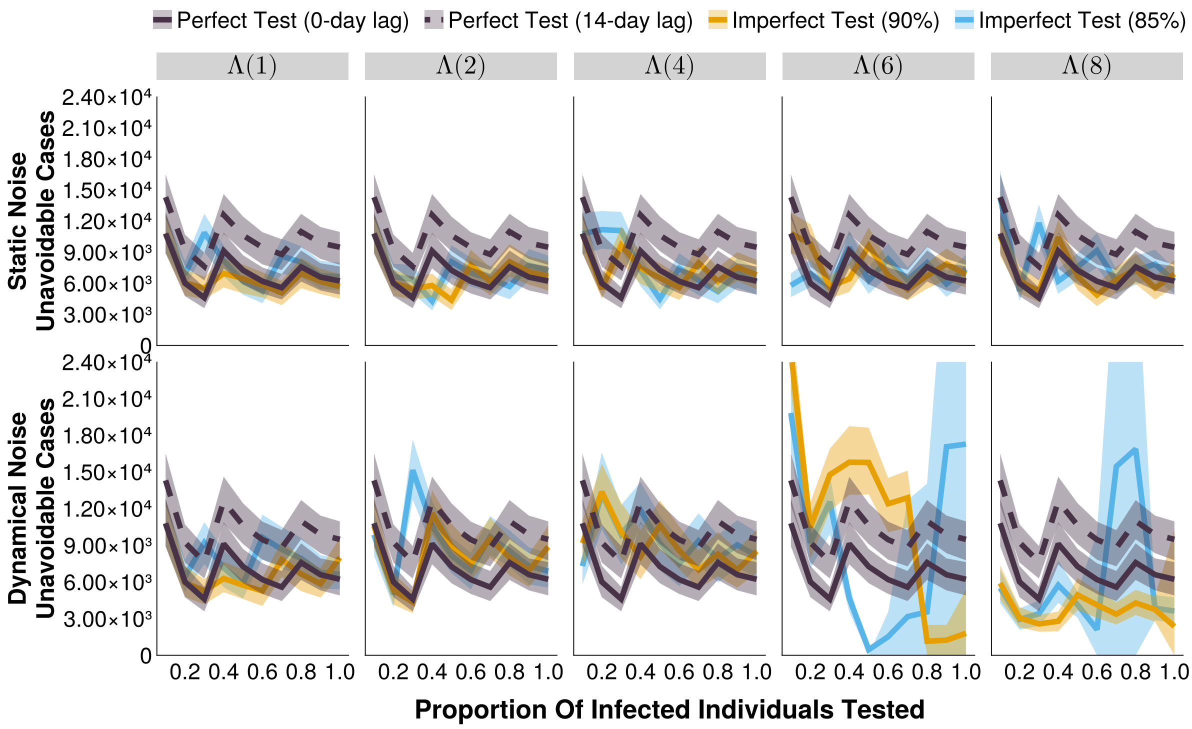


Supplemental Figure 3: The number of unavoidable cases of an outbreak detection systems under different testing rates and noise structures, at their respective optimal alert thresholds. The shaded bands illustrate the 80% central interval, and the solid/dashed lines represent the mean estimate. Each imperfect test uses the same value for both its sensitivity and specificity (either 85% or 90%). Solid lines represent tests with 0-day turnaround times, and dashed lines represent tests with result delays. $\Lambda(4)$ indicates the mean noise incidence is 4 times higher than the mean measles incidence.

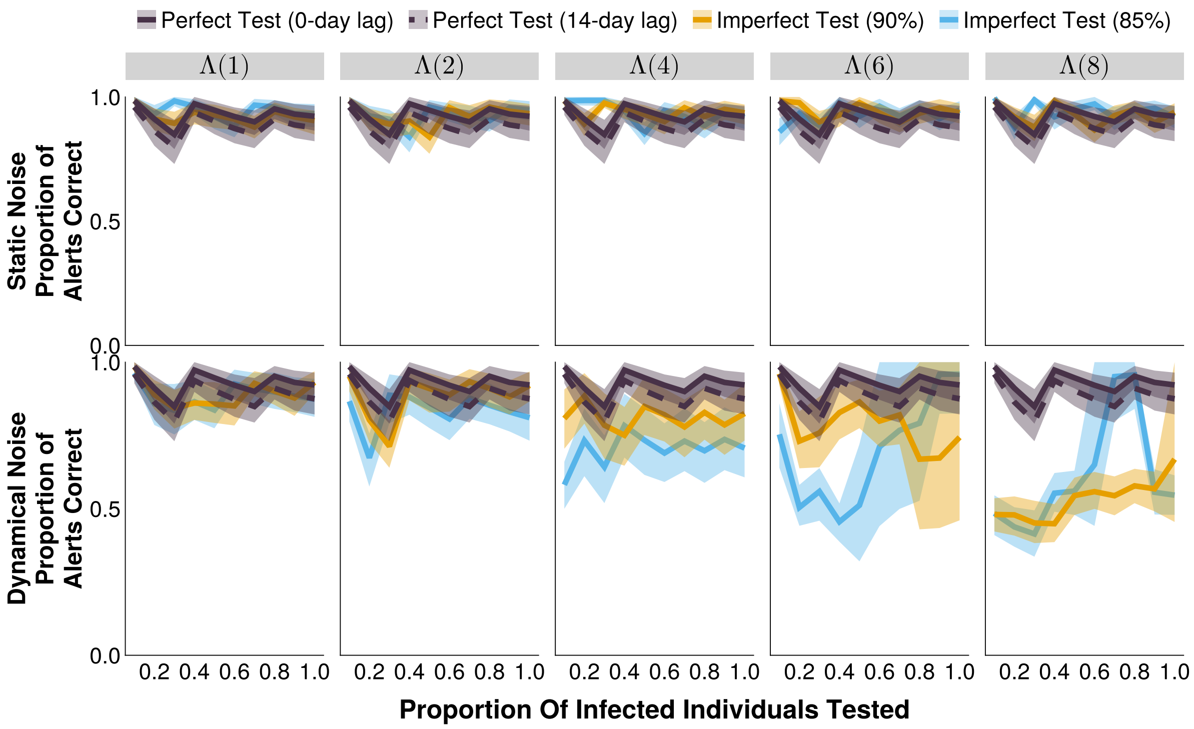


Supplemental Figure 4: The proportion of alerts of an outbreak detection system that are correctly associated with an outbreak, under different testing rates and noise structures, at their respective optimal alert thresholds. The shaded bands illustrate the 80% central interval, and the solid/dashed lines represent the mean estimate. Each imperfect test uses the same value for both its sensitivity and specificity (either 85% or 90%). Solid lines represent tests with 0-day turnaround times, and dashed lines represent tests with result delays. $\Lambda(4)$ indicates the mean noise incidence is 4 times higher than the mean measles incidence.

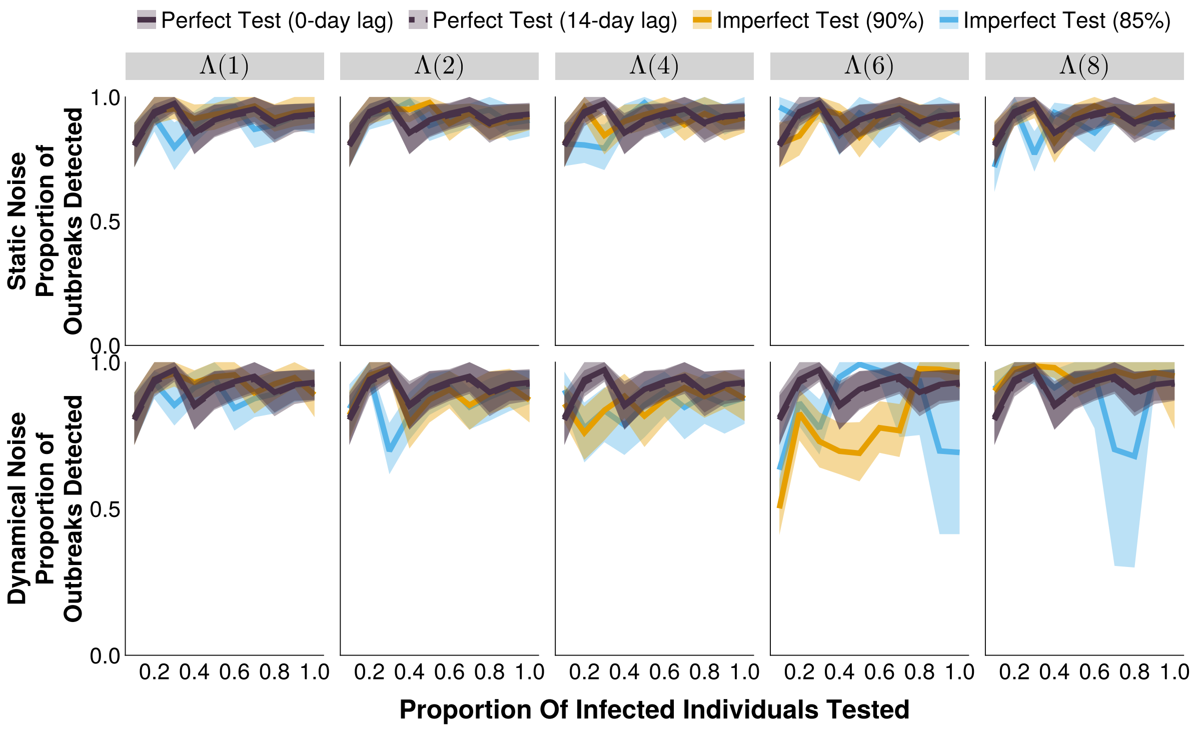


Supplemental Figure 5: The proportion of outbreaks that are correctly identified by at least one alert of an outbreak detection system, under different testing rates and noise structures, at their respective optimal alert thresholds. The shaded bands illustrate the 80% central interval, and the solid/dashed lines represent the mean estimate. Each imperfect test uses the same value for both its sensitivity and specificity (either 85% or 90%). Solid lines represent tests with 0-day turnaround times, and dashed lines represent tests with result delays. $\Lambda(4)$ indicates the mean noise incidence is 4 times higher than the mean measles incidence.

#### F-1 Score Optimizations

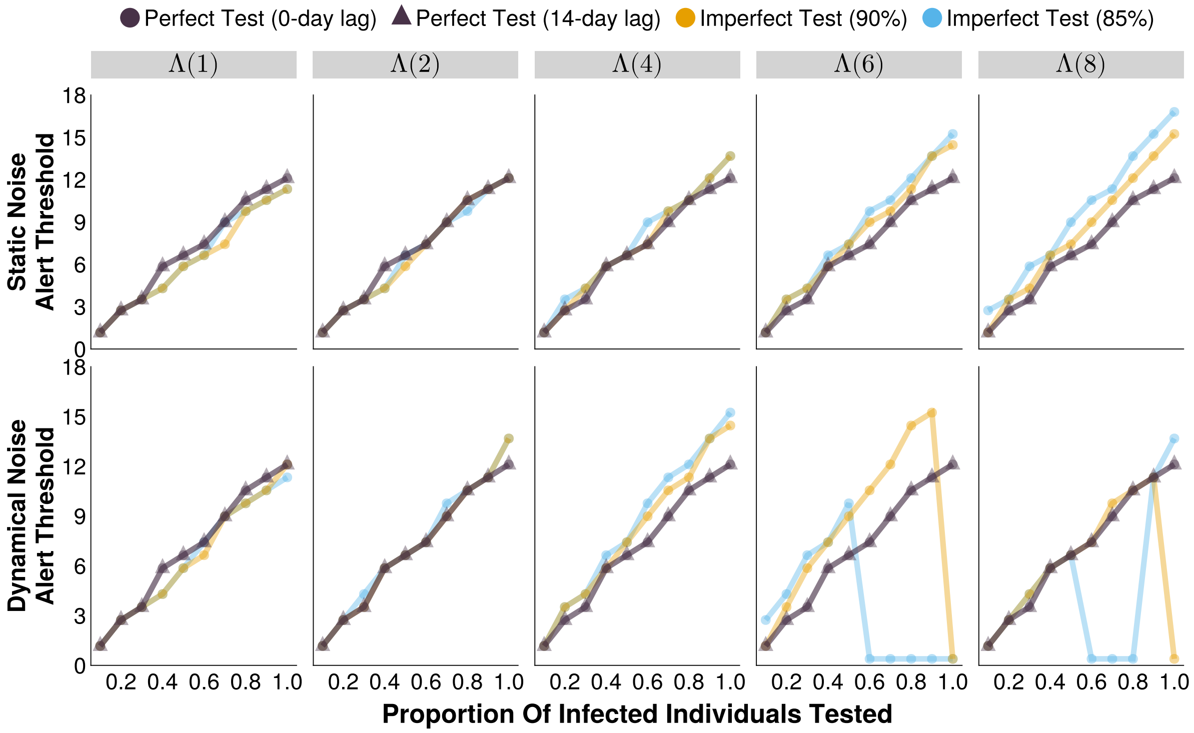


Supplemental Figure 6: The optimal alert threshold of outbreak detection systems (that maximizes outbreak detection F-1 score) under different testing rates and noise structures. Each imperfect test uses the same value for both its sensitivity and specificity (either 85% or 90%). Circular markers represent tests with 0-day turnaround times, and triangular markers represent tests with result delays. $\Lambda(4)$ indicates the mean noise incidence is 4 times higher than the mean measles incidence, for example.

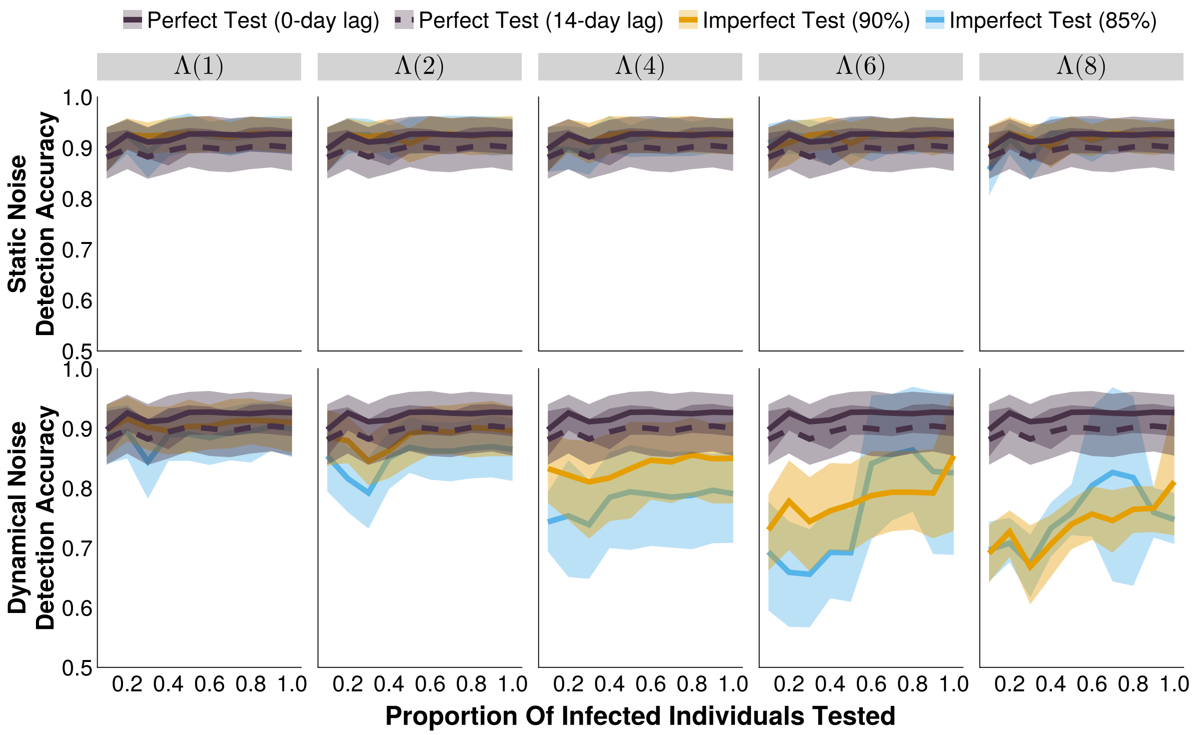


Supplemental Figure 7: The accuracy of outbreak detection systems under different testing rates and noise structures, at their respective (F-1 score) optimal alert thresholds. The shaded bands illustrate the 80% central interval, and the solid/dashed lines represent the mean estimate. Each imperfect test uses the same value for both its sensitivity and specificity (either 85% or 90%). Solid lines represent tests with 0-day turnaround times, and dashed lines represent tests with result delays. $\Lambda(4)$ indicates the mean noise incidence is 4 times higher than the mean measles incidence, for example.

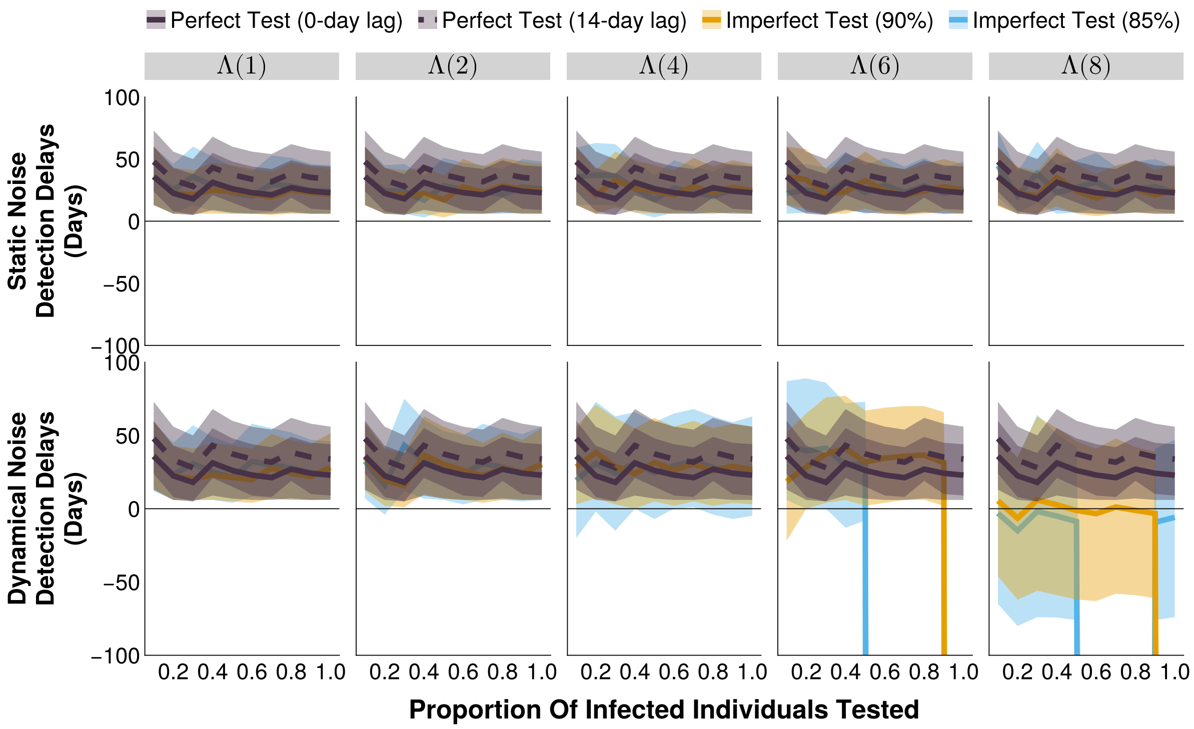


Supplemental Figure 8: The detection delay of outbreak detection systems under different testing rates and noise structures, at their respective (F-1 score) optimal alert thresholds. The shaded bands illustrate the 80% central interval, and the solid/dashed lines represent the mean estimate. Each imperfect test uses the same value for both its sensitivity and specificity (either 85% or 90%). Solid lines represent tests with 0-day turnaround times, and dashed lines represent tests with result delays. $\Lambda(4)$ indicates the mean noise incidence is 4 times higher than the mean measles incidence.

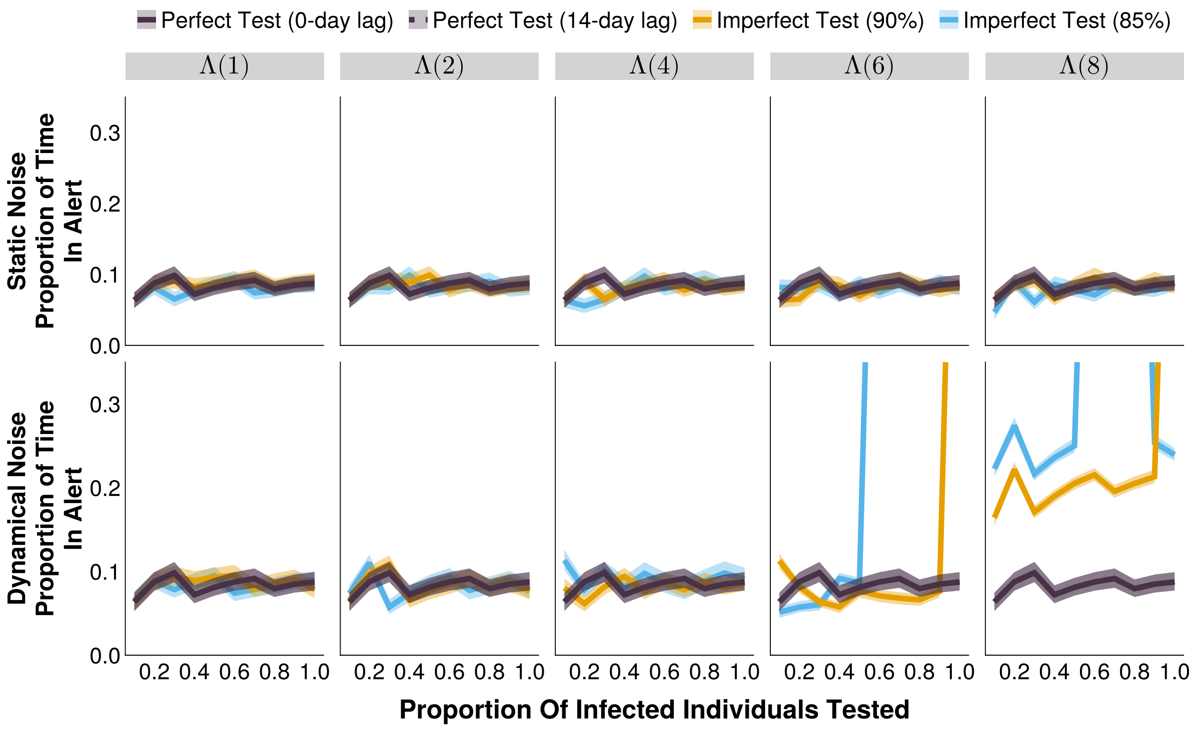


Supplemental Figure 9: The proportion of the time series in alert of outbreak detection systems under different testing rates and noise structures, at their respective (F-1 score) optimal alert thresholds. The shaded bands illustrate the 80% central interval, and the solid/dashed lines represent the mean estimate. Each imperfect test uses the same value for both its sensitivity and specificity (either 85% or 90%). Solid lines represent tests with 0-day turnaround times, and dashed lines represent tests with result delays. $\Lambda(4)$ indicates the mean noise incidence is 4 times higher than the mean measles incidence.
